# Mendelian Randomisation Analysis Suggests that Hypothyroidism Reduces Endometrial Cancer Risk

**DOI:** 10.1101/2023.07.30.23293405

**Authors:** Dylan M Glubb, Xuemin Wang, Tracy A O’Mara

**Affiliations:** Cancer Research Program, QIMR Berghofer Medical Research Institute, Brisbane, QLD 4006, Australia

**Keywords:** Endometrial cancer, thyroid dysfunction, hypothyroidism, Hashimoto’s thyroiditis, body mass index, Mendelian randomisation, genome-wide association study

## Abstract

**Background:** Thyroid dysfunction, hypothyroidism in particular, has been associated with endometrial cancer, but it remains unclear whether hypothyroidism itself or other aspects of thyroid dysfunction have a causal effect on endometrial cancer risk.

**Methods:** To clarify the effects of thyroid dysfunction phenotypes on endometrial cancer risk, we performed Mendelian randomisation analyses data from the largest available genome-wide association studies (GWAS). The robustness of associations was assessed through sensitivity analyses. To disentangle the potential influence of obesity on causal associations, we carried out multivariable Mendelian randomisation analysis.

**Results:** Mendelian randomisation analysis demonstrated a significant causal association between hypothyroidism and decreased risk of endometrial cancer (OR = 0.93; 95% CI 0.89- 0.97; p = 3.96 × 10^-4^). Hashimoto’s thyroiditis, a common cause of hypothyroidism, showed a similar, albeit nominal, association with endometrial cancer risk (OR = 0.92; 95% CI 0.86- 0.99; p = 0.03). Hypothyroidism was also significantly associated with decreased risk of endometrioid endometrial cancer (OR = 0.93; 95% CI 0.88-0.98; p = 4.02 × 10^-3^), the most common histological subtype. Sensitivity analyses confirmed the robustness of the significant associations. Multivariable Mendelian randomisation analysis revealed that BMI and hypothyroidism had independent effects on endometrial cancer risk.

**Interpretation:** This study provides evidence for a causal relationship between hypothyroidism and decreased risk of endometrial cancer. The protective effect of hypothyroidism is independent of BMI and may be related to the autoimmune effects of Hashimoto’s disease.

**Funding:** National Health and Research Council of Australia (APP1173170). Worldwide Cancer Research and Cancer Australia (22-0253).

**Research in context:** *Evidence before this study:* We searched PubMed for epidemiological and Mendelian randomisation studies containing the terms ‘endometrial cancer’ or ‘uterine cancer’ or ‘obesity’, and ‘thyroid dysfunction’ or ‘hypothyroidism’ or ‘Hashimoto’s thyroiditis’ or ‘thyroid stimulating hormone’ or ‘triiodothyronine’ or ‘thyroxine’ or ‘thyroid peroxidase’ or ‘Graves’ disease’ or ‘hyperthyroidism’ without date restrictions. These searches revealed that thyroid dysfunction has been a subject of interest in relation to endometrial cancer. Indeed, observational studies have previously suggested an association between hypothyroidism and increased risk of endometrial cancer although the nature of these studies have limited their ability to establish causal relationships. Additionally, the potential confounding effect of obesity, a shared risk factor for both endometrial cancer and hypothyroidism, further complicates these relationships.

*Added value of this study:* By employing Mendelian randomization analysis, a powerful approach that reduces confounding, we identified a robust causal association between hypothyroidism and a decreased risk of endometrial cancer. This finding challenges the previously suggested association between hypothyroidism and increased endometrial cancer risk. Notably, our study did not find evidence that thyroid hormone levels influence endometrial cancer risk. However, we observed a suggestive association between Hashimoto’s thyroiditis, a common cause of hypothyroidism. Furthermore, we demonstrated the independent effects of body mass index (a surrogate measure for obesity) and hypothyroidism on endometrial cancer risk, with hypothyroidism potentially attenuating the impact of body mass index.

*Implications of the available evidence:* The identification of hypothyroidism as a protective factor for endometrial cancer raises intriguing questions about the disease’s pathogenesis. The available evidence suggests involvement of autoimmune effects, highlighting the need for further studies investigating the role of immune responses in endometrial cancer development. By elucidating the specific pathways and molecular mechanisms underlying the relationship of endometrial cancer with hypothyroidism, we may uncover potential targets for preventive or therapeutic interventions.

## Introduction

Endometrial cancer is the most common gynecological cancer in developed countries, with an increasing incidence that appears to be related to obesity.^1^ Indeed, epidemiological studies have shown that obesity is one of the strongest risk factors for endometrial cancer (reviewed by Crosbie *et al.*^2^). To establish a causal relationship between obesity and endometrial cancer risk, Mendelian randomisation analyses have been conducted using germline genetic variants strongly associated with body mass index (BMI) as instrumental variables (IVs) (reviewed by Wang *et al.*^3^). By leveraging the random assortment of germline variants, which remain unaffected by disease or environmental factors, the Mendelian randomisation approach enables the assessment of causality while mitigating biases arising from confounding and reverse causation.^4^ As a result, Mendelian randomisation analysis has increasingly been used to assess relationships been risk factors and disease, and has successfully confirmed the causality of other endometrial cancer risk factors, including levels of sex hormones such as estradiol and testosterone (reviewed by Wang *et al.*^3^).

The thyroid gland is an important endocrine organ that in response to thyroid stimulating hormone (TSH) produces two main hormones: triiodothyronine (T3) and thyroxine (T4). These hormones have multiple functions relevant to endometrial cancer development. For example, thyroid hormones regulate appetite, lipid storage and metabolic rate,^5^ and also affect the morphophysiology of the endometrium.^6^ Thyroid dysfunction has also been linked to endometrial cancer, with a prior diagnosis of thyroid disease associated with increased endometrial cancer risk in a Danish record linkage study.^7^ Furthermore, uterine cancer cases, which primarily consist of endometrial cancer, have been found to be more prevalent among Italian patients with thyroid disease, compared to the general population.^8^ Thyroid dysfunction can manifest as hypothyroidism, diagnosed by high TSH levels and low T4 levels, or, conversely, as hyperthyroidism, diagnosed by low TSH levels and high T3 and T4 levels. Notably, hypothyroidism is a common comorbidity for endometrial cancer (reviewed by Wang *et al.*^9^) and, consistent with this observation, increased TSH levels have been found in endometrial cancer patients in a small case-control study.^10^

Another factor to consider in the relationship between thyroid dysfunction and endometrial cancer is obesity. Hypothyroidism and treatment of hyperthyroidism have been associated with weight gain or obesity (reviewed by Laurberg *et al.*^11^). However, Mendelian randomisation analyses have not provided evidence that alterations in levels of TSH, T3 or T4 affect obesity ^12^. Intriguingly, both epidemiological and Mendelian randomisation studies suggest that obesity is a risk factor for thyroid dysfunction,^12^ hypothyroidism in particular.^13,14^

Mendelian randomisation analyses have been performed to assess the effects of thyroid dysfunction on uterine cancer using genetic data from a limited number (n = 1,931) of cases from the UK Biobank, primarily composed of endometrial cancer patients.^15^ No significant effects were observed for hypothyroidism, hyperthyroidism or T4 but TSH was found to be nominally associated with reduced cancer risk, inconsistent with the previous observational findings. In the current study, we aimed to provide clarity on the effects of thyroid dysfunction by using genetic data from a much larger number of endometrial cancer cases (n = 12,270) provided by the Endometrial Cancer Association Consortium (ECAC).^16^ These Mendelian randomisation analyses included genetic instruments for hypothyroidism, hyperthyroidism, TSH, T3 and T4. Considering that autoimmune thyroid disease, including Hashimoto’s thyroiditis (hypothyroidism) and Graves’ disease (hyperthyroidism), is responsible for ∼90% of all thyroid diseases,^17^ we included these conditions in our analyses; in addition to thyroid peroxidase (TPO) antibody positivity, a marker for autoimmune thyroid disease. Furthermore, given the role of obesity in the development of thyroid dysfunction and endometrial cancer, we have attempted to disentangle independent causal effects using multivariable Mendelian randomisation analysis.

## Methods

### Genome-wide association study (GWAS) summary statistics

For Mendelian randomisation analyses, we sourced GWAS summary statistics from studies described in **Supplementary Table 1**. For endometrial cancer risk, data were derived from the largest GWAS meta-analysis of endometrial cancer risk.^16^ To avoid potential bias from overlapping samples with other GWAS datasets used in the current study, UK Biobank samples had been removed, resulting in 12,270 endometrial cancer cases and 46,126 controls for the generation of GWAS summary statistics.^18^ For secondary analyses using this dataset, we used summary statistics from GWAS of cases with either endometrioid (8,758 cases and 46,126 controls) or non-endometrioid histology (1,230 cases and 35,447 controls). Histological subtypes of endometrial cancer were confirmed based on pathology reports, and details have been described previously.^16,19^

For autoimmune thyroid disease, we used GWAS summary statistics from UK Biobank and Iceland cases and controls.^20^ Cases were defined as individuals who had received a diagnosis of Graves’ disease, Hashimoto’s thyroiditis, or hypothyroidism, or had received levothyroxine treatment; individuals with known non-autoimmune causes of hypothyroidism (i.e. thyroid cancer and drug-induced hypothyroidism) were excluded.^20^ GWAS summary statistics for hypothyroidism were derived from a meta-analysis of cases and controls of European ancestry from UK Biobank and FinnGen, with cases identified based on clinical diagnosis of hypothyroidism or treatment with levothyroxine.^21^ GWAS summary statistics for hyperthyroidism were obtained from a meta-analysis of cases and controls with European ancestry from 19 cohorts.^22^ For Graves’ disease and Hashimoto’s thyroiditis, GWAS had been performed using participants with European ancestry but included only 2,285 and 462 cases, respectively.^23^ As these GWAS had limited power to detect meaningful associations for these diseases, we used GWAS summary statistics from meta-analyses of two European and one East Asian populations.^24^ For Graves’ disease, 1,678 cases from UKBB and FinnGen (release 3) were meta-analysed with 2,809 cases from Biobank Japan; for Hashimoto’s thyroiditis, 15,654 cases from UK Biobank and FinnGen were meta-analysed with 537 cases from Biobank Japan.

In addition to disease-specific GWAS, we also used summary statistics for thyroid hormones or markers. GWAS for free T3 levels had been performed in disease-free participants from three Croatian cohorts but full summary statistics were not available.^25^ For TPO antibody positivity, we used data from a two-stage GWAS of 11 European populations but full summary statistics were also not available.^26^ For both free T4 and TSH levels, female-stratified and sex-combined GWAS summary statistics were used. For female-stratified and sex-combined free T4, GWAS summary statistics were derived from a meta-analysis of 26 cohorts.^27^ For female TSH levels, summary statistics were available from a GWAS meta-analysis of women from 30 cohorts,^27^ and for sex-combined TSH levels, we used summary statistics from a GWAS meta-analysis of three studies.^28^ For BMI, we used GWAS summary statistics derived from 434,794 women of European ancestry in a meta-analysis of the Genetic Investigation of Anthropometric Traits consortium and UK Biobank.^29^

### IV selection

GWAS summary statistics from all phenotypes were used to identify genetic variants associated with exposures at genome-wide significance levels (p < 5 × 10^-8^); no such variants were available for non-endometrioid endometrial cancer. To identify independent IVs, we used a window of 10 Mb and maximal linkage disequilibrium of r^2^=0.001 between instruments in PLINK.^30^ The linkage disequilibrium reference used a random sample of 10,000 unrelated participants from the UK Biobank.^18^ Palindromic variants (i.e. those with A/T or G/C alleles) with a minor allele frequency ≥ 0.42 were excluded to prevent errors due to strand ambiguity. Variants from the *SH2B3* locus were also excluded as this is a pleiotropic locus and, through GWAS, has been associated with many traits including hypothyroidism,^31^ Hashimoto’s thyroiditis,^24^ and endometrial cancer.^16^ To assess weak instrument bias, we calculated the *F*-statistic^32^ for each instrumental variable and all IVs of an exposure using the formula *F* = R^2^ × (N-1-k) / ((1-R^2^) × k), where R^2^ is the proportion of variance explained by the instrumental variable, N is the sample size, and k is the number of IVs included in the analysis.^33^ For the calculation of *F*-statistic for each instrumental variable, k equals to 1. The proportion of variance explained by IVs was calculated using the formula in the Supplementary Methods of Yarmolinsky *et al.*^33^ An *F*-statistic > 10 is often used to indicate sufficient instrument strength. For quantitative traits, such as circulating thyroid hormones (i.e., TSH, T3, or T4), IVs are reported as the association with one standard deviation increase in the levels of the respective hormone. Details of IVs used in Mendelian randomisation analyses can be found in **Supplementary Table 2**.

### Mendelian randomisation analysis

To assess causal relationships, we used the two sample Mendelian randomisation framework,^34^ following the Strengthening the Reporting of Observational Studies in Epidemiology using Mendelian Randomisation (STROBE-MR) checklist (**Supplementary Information**).^35,36^ For free T3, the only exposure with a single IV, we used the Wald ratio to estimate effects. For all other exposures, multiple IVs were combined using the inverse-variance weighted (IVW) random effect method as the primary analysis. The IVW method uses all available IVs and thus has the most power to detect an association, assuming that the IVs meet certain criteria: (1) they are reliably associated with the exposure of interest; (2) they are not associated with any confounders that mediate the association between exposure and outcome; and (3) they only affect the outcome through the exposure, without any horizontal pleiotropy. To detect violations in the underlying Mendelian randomisation assumptions, we assessed heterogeneity in the causal effect estimates across IVs by calculating Cochran’s Q statistic and its associated p-value.^37^ Wald’s ratio, IVW and Cochran’s Q statistic analyses were implemented in the TwoSampleMR (version 0.5.6) R package.^38^

We used the online Mendelian randomisation power calculator mRnd (https://shiny.cnsgenomics.com/mRnd/)^39^ to calculate the power for Mendelian randomisation analyses to detect an association between one standard deviation change in the trait of interest under three scenarios reflecting weak, moderate, and strong effects with cut-offs set to OR > 1.1, OR > 1.2, and OR > 1.4, respectively. For the calculation of power for continuous outcomes, variances of the exposure and outcome variables were both set to 1 given GWAS for outcome were conducted on the inverse-normalised variable. Results of the power calculations can be found in **Supplementary Table 3**.

To determine the effects of thyroid dysfunction phenotypes on endometrial cancer risk, we used a Bonferroni correction threshold for significance, adjusting for the testing of 11 traits (p < 4.5 × 10^-3^). We also performed bidirectional analyses, using thyroid dysfunction phenotypes as outcomes and risk of endometrial cancer (and its histological subtypes) as exposures. To investigate the involvement of BMI in potential causal relationships, we conducted bidirectional MR analysis between BMI and exposures that were significantly associated with endometrial cancer risk. We were unable to study free T3 or TPO antibody positivity as outcomes due to the unavailability of full GWAS summary statistics for these phenotypes. We were also unable to examine the effects of non-endometrioid endometrial cancer risk as an exposure due to the absence of IVs for this trait.

### Mendelian randomisation sensitivity analyses

IV assumptions (2) and (3) are often violated in a Mendelian randomisation setting. Therefore, to account for potential violations of these assumptions and evaluate the robustness of significant associations identified by IVW analysis, we conducted sensitivity analyses using the MR-Egger, weighted median and the Mendelian Randomisation pleiotropy residual sum and outlier (MR-PRESSO) methods. The MR-Egger method relaxes assumption (2) by allowing a non-zero regression intercept.^40^ This provides an unbiased causal estimate even when assumption (2) is violated for all IVs, as long as the magnitude of the horizontal pleiotropic effects is independent of the variant-exposure effects. The MR-Egger method also provides a formal test of the existence of horizontal pleiotropy.^40^ The weighted median method assumes at least half of the variants are valid IVs and so provides a reliable estimate even if the assumptions for some IVs are violated. ^41^ Lastly, MR-PRESSO identifies and adjusts for outlier IVs displaying horizontal pleiotropic effects.^42^ As these sensitivity analyses do not yield reliable results when using small numbers of IVs, they were only performed when a minimum of eight IVs were available. The MR-Egger, and weighted median methods were implemented in the TwoSampleMR (version 0.5.6) R package;^38^ additionally, we used the MR-PRESSO (version 1.0) R package. A significance threshold of p < 0.05 was applied for sensitivity analyses.

### Multivariable Mendelian randomisation analysis

To investigate the independent effects of BMI and hypothyroidism on endometrial cancer risk, we conducted multivariable Mendelian randomisation analysis. We first clumped genetic variants with GWAS summary statistics for BMI and hypothyroidism together in PLINK using a window of 10 Mb and a maximal linkage disequilibrium of r^2^=0.001 between variants, as per the univariable Mendelian randomisation analyses. As there are far more genetic variants associated with BMI, we preferentially selected variants for hypothyroidism that were in linkage disequilibrium with variants associated with BMI to maximise the number of IVs for hypothyroidism. A list of the selected IVs are provided in **Supplementary Table 4**. The multivariable analysis was performed using the MendelianRandomisation (version 0.6.0) R package.^43^ R code for the multivariable analysis can be found in **Supplementary Information**. To confirm the robustness of the results from the multivariable analysis, we also reran univariable Mendelian randomisation analyses with endometrial cancer risk as the outcome using only IVs included the multivariable analysis for BMI or hypothyroidism risk.

### Ethics

The analyses were based on publicly available summary-level data that have been approved by relevant review boards and participants have provided informed consent.

## Results

### Hypothyroidism is causally associated with decreased endometrial cancer risk

Mendelian Randomisation analysis revealed a significant causal association between hypothyroidism and decreased endometrial cancer risk (OR = 0.93; 95% CI 0.89-0.97; p = 3.96 × 10^-4^; **Figure 1**). Although no other thyroid dysfunction phenotype passed the Bonferroni multiple correction threshold for association with endometrial cancer risk (p < 4.5 × 10^-3^; **Figure 1**), we did observe a nominally significant association between Hashimoto’s thyroiditis and decreased endometrial cancer risk (OR = 0.92, 95% CI 0.86-0.99; p = 0.03; **Figure 1**). As Hashimoto’s thyroiditis accounts for most cases of hypothyroidism in developed countries,^44^ the two findings are consistent with each other. However, the power of our Mendelian randomisation analyses was much greater for studying the effects of hypothyroidism than Hashimoto’s thyroiditis (**Supplementary Table 3**), potentially explaining the difference in statistical significance. In analysis of endometrial cancer histological subtypes, we also found a significant causal association between hypothyroidism and decreased endometrioid endometrial cancer risk (OR = 0.93, 95% CI 0.88-0.98; p = 4.02 × 10^-3^; **Figure 1**). However, there was some evidence of IV heterogeneity in this analysis from Cochran’s Q statistic (p < 0.05; **Supplementary Table 5**). Bidirectional Mendelian randomisation analysis did not support effects of endometrial cancer risk on thyroid dysfunction phenotypes (**Supplementary Table 6**).

**Figure 1.**
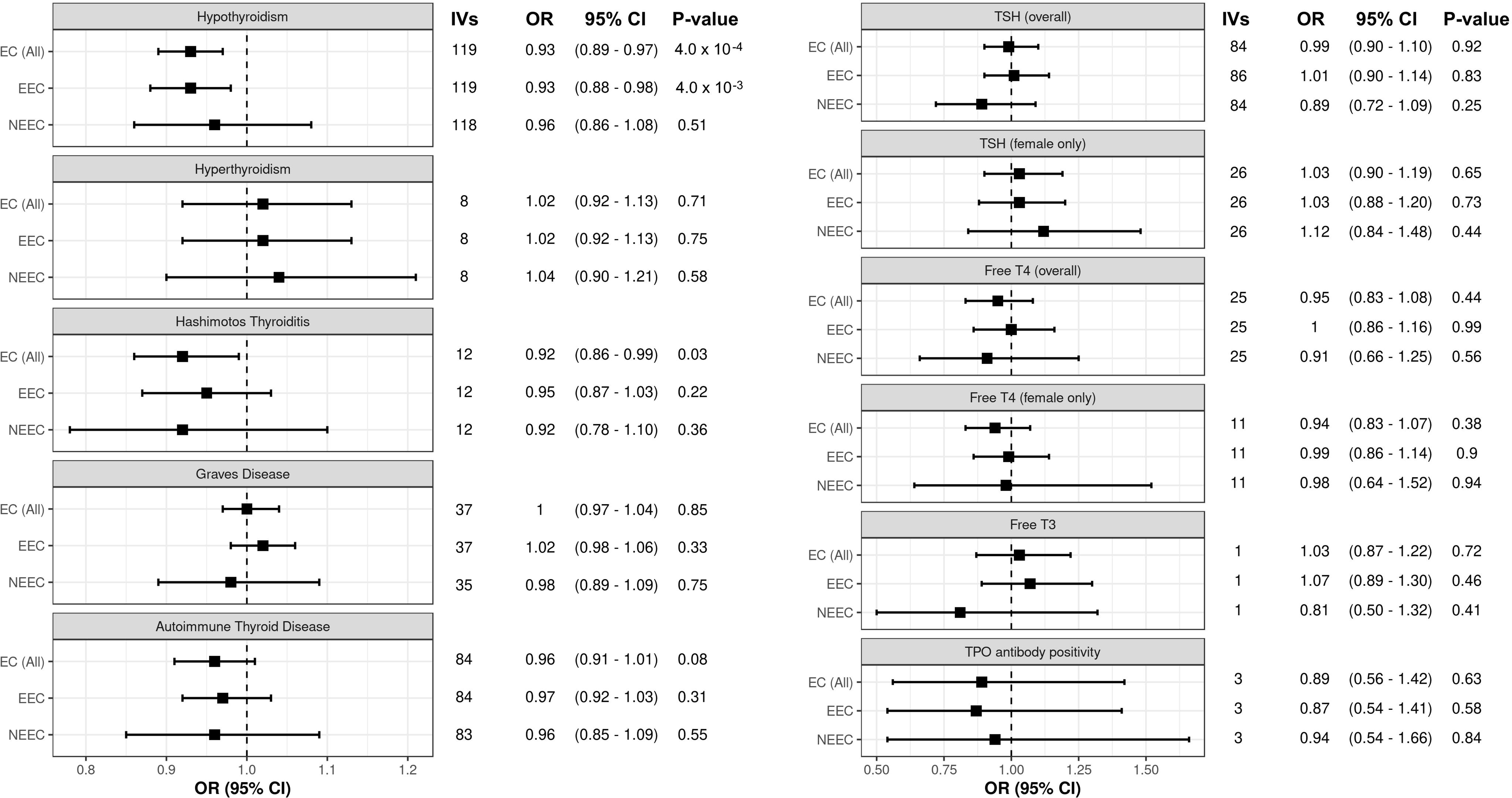
Mendelian Randomisation analysis of the effect of thyroid dysfunction phenotypes on endometrial cancer risk. Each panel displays the inverse-variance weighted (IVW) analysis results for the associations between thyroid dysfunction phenotypes (as indicated) and the risk of all endometrial cancer (EC (All)), endometrioid endometrial cancer (EEC), and non-endometrioid endometrial cancer (NEEC). Numbers of instrumental variables (IVs) used in each analysis are shown.

### Sensitivity analysis of associations between thyroid dysfunction and endometrial cancer risk

We performed sensitivity analyses to evaluate the associations of hypothyroidism and Hashimoto’s disease with endometrial cancer risk. These analyses supported the association between hypothyroidism and decreased risk of endometrial cancer, as demonstrated by significant associations in the weighted median and MR-PRESSO analyses (**Figure 2**). For the association of hypothyroidism with endometrioid endometrial cancer risk, the MR-Egger analysis showed a discordant direction of effect (**Figure 2**) and suggested the presence of horizontal pleiotropy (**Supplementary Table 5**). However, by removing an IV that contributed to heterogeneity (**Supplementary Table 5**), the MR-PRESSO analysis demonstrated a significant association between hypothyroidism and decreased endometrioid endometrial cancer risk, consistent with the IVW analysis (**Figure 2**). Lastly, the MR-Egger analysis again provided a discordant direction of effect for the association of Hashimoto’s thyroiditis with endometrial cancer risk (**Figure 2**), but there was no evidence of pleiotropy (**Supplementary Table 5**). Furthermore, the directionality of the MR-PRESSO and weighted median analyses were consistent with the association of Hashimoto’s thyroiditis with reduced endometrial cancer risk and (**Figure 2**).

**Figure 2.**
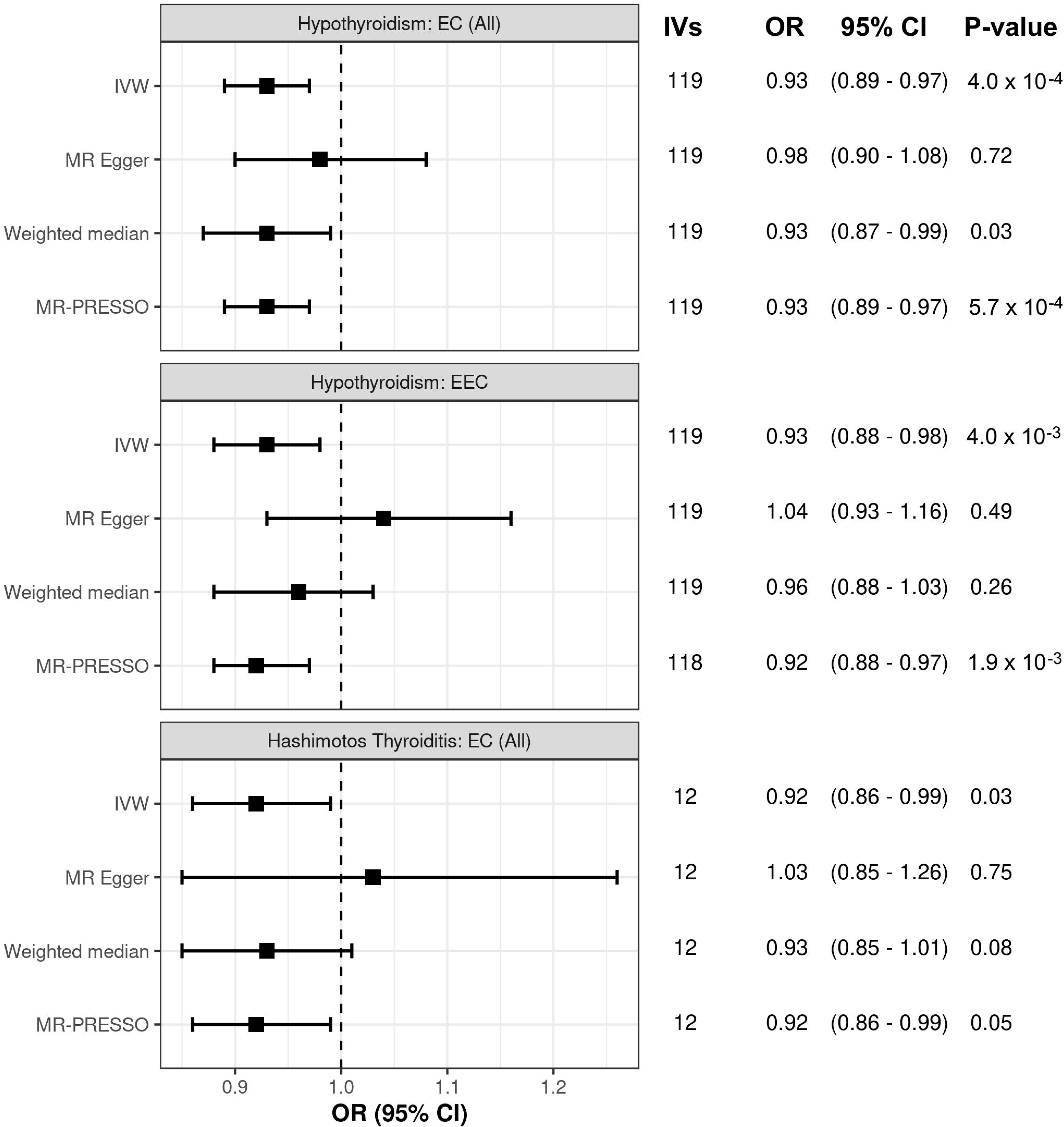
Mendelian Randomisation sensitivity analyses for associations of hypothyroidism and Hashimoto’s thyroiditis with endometrial cancer risk. Sensitivity analysis of the association between hypothyroidism and risk of all endometrial cancer (EC (All)), hypothyroidism and risk of endometrioid endometrial cancer (EEC), and between Hashimoto’s thyroiditis and risk of EC (All). Panels display sensitivity analyses using MR Egger, weighted median and MR-PRESSO approaches, and the primary inverse-variance weighted (IVW) analysis. Numbers of instrumental variables (IVs) used in each analysis are shown.

### BMI and hypothyroidism are independently associated with endometrial cancer risk

To assess the effect of BMI on the relationship between hypothyroidism and endometrial cancer risk, we firstly performed bidirectional Mendelian randomisation analysis of BMI and hypothyroidism. Our findings revealed that increased BMI was very strongly associated with increased hypothyroidism risk but there was no evidence for a significant effect of predisposition to hypothyroidism on BMI (**Supplementary Table 7**). Although some IV heterogeneity was detected in the IVW analysis of BMI’s effect on hypothyroidism, the finding remained significant in all sensitivity analyses (**Supplementary Table 7**). We next performed multivariable Mendelian randomisation, which demonstrated that both BMI and hypothyroidism independently associated with endometrial cancer risk (**Figure 3A-B**). Similar associations were also observed for endometrioid endometrial cancer risk (**Figure 3A-B**). Notably, the multivariable analyses showed slightly stronger associations compared to the univariable analyses, particularly for the effect of BMI on endometrial cancer risk, including the histological subtypes (**Figure 3B**). These observations suggest that hypothyroidism acts as a mediator in the causal pathway between BMI and endometrial cancer (**Figure 3C**). Specifically, increased BMI raises the risk of hypothyroidism, which in turn has a protective effect on endometrial cancer.

**Figure 3.**
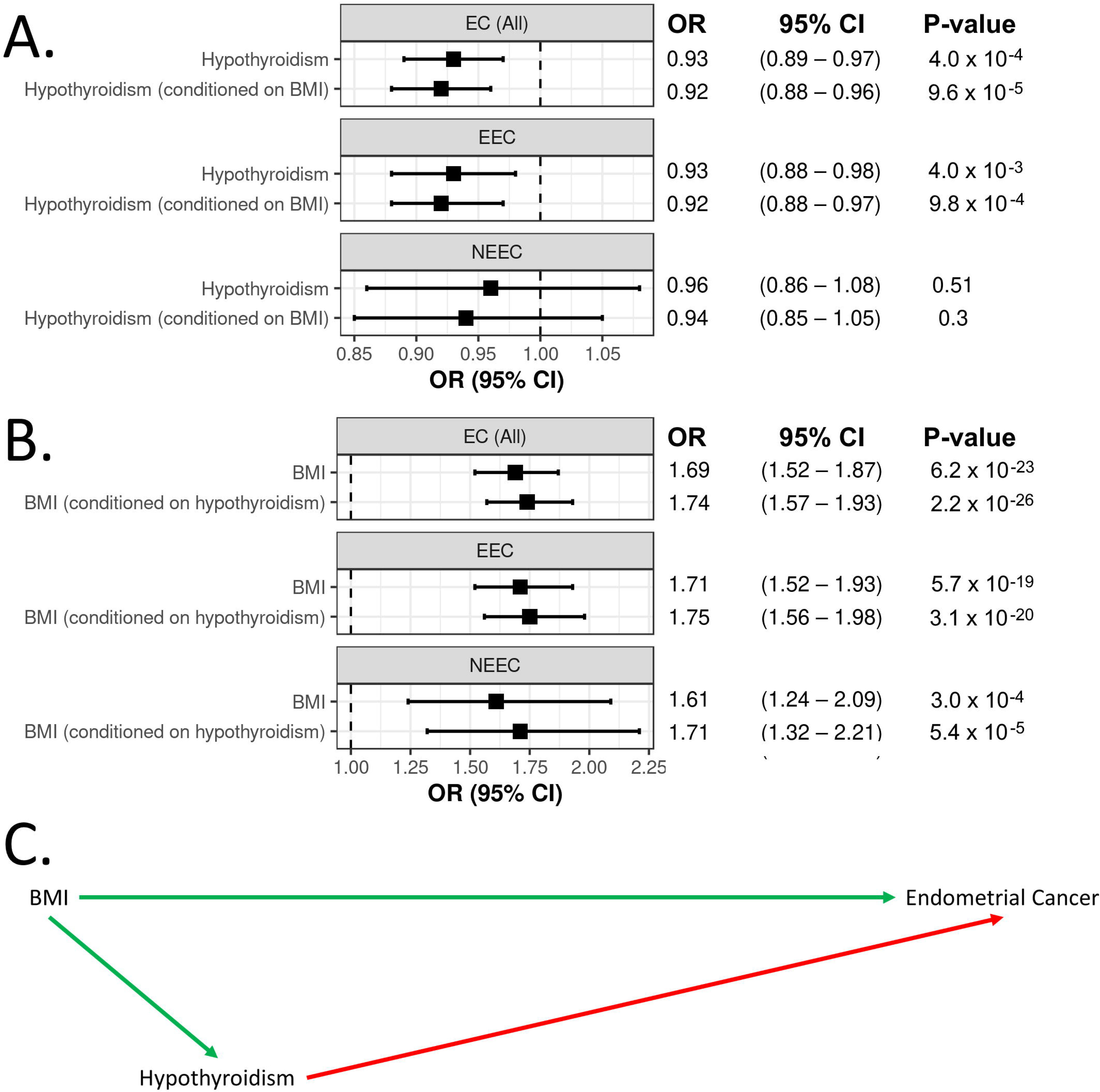
Multivariable Mendelian Randomisation analysis of the effects of BMI and hypothyroidism on endometrial cancer risk. Panels **A** and **B** show the univariable or multivariable associations of BMI and hypothyroidism, respectively, with risk of all endometrial cancer (EC (All)), endometrioid endometrial cancer (EEC), and non-endometrioid endometrial cancer (NEEC). The relationships between BMI, hypothyroidism and endometrial cancer are depicted in a directed acyclic graph (**C**). Positive associations are indicated by green arrows, while negative associations are shown by red arrows.

## Discussion

Mendelian Randomisation analysis revealed a significant causal association between hypothyroidism and decreased endometrial cancer risk. Additionally, we found a suggestive association between Hashimoto’s disease and reduced endometrial cancer risk. This finding indicates that the effect of hypothyroidism on endometrial cancer risk may be driven by Hashimoto’s thyroiditis, the leading cause of hypothyroidism in developed countries. Subtype analysis uncovered a significant causal association between hypothyroidism and decreased risk of endometrioid endometrial cancer, which constitutes the majority of endometrial cancer cases both in the general population and the endometrial cancer GWAS dataset. Sensitivity analyses demonstrated the robustness of the associations of hypothyroidism with endometrial cancer risk and provided support for the potential effect of Hashimoto’s thyroiditis.

We did not detect any significant associations between thyroid hormone levels and endometrial cancer risk in our analysis. This contrasts with a previous Mendelian randomisation study by Yuan *et al.* which reported a nominal association between TSH levels and reduced uterine cancer risk.^15^ Yuan *et al.* also studied 21 other site-specific cancers and found that TSH levels were associated with reduced risk of overall cancer, with individual associations for reduced risk of breast and thyroid cancer. Despite having sufficient statistical power to detect weak to moderate effects of TSH levels, we found no evidence of such an association with endometrial cancer risk.

The mechanism underlying the potential protective effect of hypothyroidism on endometrial cancer remains unclear. It is noteworthy that hypothyroidism often develops in endometrial cancer patients as a result of immunotherapy,^45,46^ suggesting that the effect of hypothyroidism on endometrial cancer may be linked to autoimmunity, driven by Hashimoto’s disease. This hypothesis is further supported by robust associations between systemic lupus erythematosus (SLE), another autoimmune disease, and reduced endometrial cancer risk in Mendelian randomisation analyses.^47^ Furthermore, Mendelian randomisation analysis indicates that there is bidirectional relationship between hypothyroidism and SLE, with a particularly strong effect of hypothyroidism on increased SLE risk.^48^ The protective effect of SLE on endometrial cancer has been proposed to be related to antinuclear autoantibodies that target cancer cells with defective DNA repair,^47^ potentially indicating similar effects for thyroid autoantibodies. Supporting this hypothesis, an observational study found that uterine cancer was significantly less prevalent in thyroid disease patients who had thyroid autoantibodies compared to thyroid disease patients who tested negative for autoantibodies.^8^ These observations highlight the need for further studies investigating the role of autoimmune responses in endometrial cancer development. Indeed, understanding the specific pathways and molecular mechanisms underlying the relationship between endometrial cancer and hypothyroidism may offer new avenues for preventive or therapeutic strategies.

To evaluate the impact of BMI on the relationship between hypothyroidism and endometrial cancer risk, we conducted additional Mendelian Randomisation analyses. Bidirectional analysis revealed a strong association between increased BMI and hypothyroidism risk, while there was no significant effect of hypothyroidism on BMI. In multivariable analyses, both BMI and hypothyroidism demonstrated independent effects on endometrial cancer (including the endometrioid subtype): hypothyroidism remained associated with decreased risk, while BMI was associated with increased risk. Notably, we found that adjusting for hypothyroidism slightly strengthened the association between BMI and endometrial cancer risk. Thus, hypothyroidism appears be a mediator of the relationship, attenuating the effect of BMI on endometrial cancer risk. Moreover, this finding provides an explanation for the association of hypothyroidism with endometrial cancer in observational studies. Specifically, as obesity is an endometrial cancer risk factor and is more prevalent among endometrial cancer patients, it follows that the prevalence of hypothyroidism will also be increased in these individuals.

Several limitations should be acknowledged when interpreting our results. Firstly, the statistical power of our analyses varied across thyroid dysfunction phenotypes, with limited power to detect low to moderate effects of phenotypes such as hyperthyroidism, Graves’ disease, and TPO antibody positivity. As a result, the lack of significant associations for these phenotypes could be the result of insufficient statistical power rather than a true absence of association. This limitation highlights the need for larger GWAS datasets and additional IVs to capture more of the trait variance for future analyses. Secondly, the analyses of the non-endometrioid subtype were exploratory in nature and should also be interpreted with caution. There was low statistical power to detect associations with this outcome and significant heterogeneity exists within the non-endometrioid subtype, which encompasses tumours with serous, clear cell, and other histologies. Future GWAS with larger sample sizes and more comprehensive characterization of non-endometrioid cases are warranted to better understand the associations within this subtype. Finally, the generalisability of our findings may be limited to populations of European ancestry, as the genetic data used in our study predominantly represent individuals from this genetic background. To validate and generalise these findings, it is crucial for future studies to encompass more diverse populations.

In summary, our study provides clarity into the relationship between thyroid dysfunction and endometrial cancer risk. Mendelian Randomisation analyses provide evidence of a causal association between hypothyroidism and decreased risk of endometrial cancer, particularly the endometrioid subtype. Notably, we did not find any significant associations between thyroid hormone levels and endometrial cancer risk. The underlying mechanism for the potential protective effect of hypothyroidism is unclear and further study is needed, but autoimmunity, driven by Hashimoto’s disease, may play a role. Multivariable analyses revealed that BMI and hypothyroidism independently influence endometrial cancer risk, with hypothyroidism appearing to act as a mediator and attenuating the effect of BMI. Future investigations should use larger GWAS datasets, when available, to clarify the lack of association for other phenotypes, and consider diverse populations to enhance the generalisability of the findings.

## Contributors

D.G., X.W. and T.A.O conceived and designed the study. X.W. performed the data analyses. T.A.O supervised the study and data analyses. D.G. and T.A.O interpreted the findings of the data analyses. D.G. and X.W. drafted the manuscript. All authors critically revised the manuscript for intellectual content.

## Supporting information

STROBE-MR

## Data Availability

All data produced in the present work are contained in the manuscript

https://www.ebi.ac.uk/gwas/

## Acknowledgements

This research was funded by an Investigator grant from the National Health and Research Council of Australia, grant number APP1173170, and a project grant co-funded by Worldwide Cancer Research and Cancer Australia, grant number 22-0253.

## Data Sharing Statement

All data analysed in this study are publicly available and their sources have been referenced throughout the manuscript and supplementary materials.

