## Supplementary material for "Mendelian Randomisation Analysis Suggests that Hypothyroidism Reduces Endometrial Cancer Risk": STROBE-MR

**STROBE-MR checklist of recommended items to address in reports of Mendelian randomization studies**^1^ ^2^

| **Item No.** | **Section** | **Checklist item** | **Page No.** | **Relevant text from manuscript** |
| --- | --- | --- | --- | --- |
| 1 | **TITLE and ABSTRACT** | Indicate Mendelian randomization (MR) as the study’s design in the title and/or the abstract if that is a main purpose of the study | 1-2 | “Mendelian Randomisation Analysis Suggests that Hypothyroidism Reduces Endometrial Cancer Risk” |
|  | **INTRODUCTION** |  |  |  |
| 2 | **Background** | Explain the scientific background and rationale for the reported study. What is the exposure? Is a potential causal relationship between exposure and outcome plausible? Justify why MR is a helpful method to address the study question | 5-6 | “Thyroid dysfunction has also been linked to endometrial cancer, with a prior diagnosis of thyroid disease associated with increased endometrial cancer risk in a Danish record linkage study [7]. Furthermore, uterine cancer cases, which primarily consist of endometrial cancer, have been found to be more prevalent among Italian patients with thyroid disease, compared to the general population [8]... Notably, hypothyroidism is a common comorbidity for endometrial cancer (reviewed in [9]) and, consistent with this observation, increased TSH levels have been found in endometrial cancer patients in a small case-control study [10].”  “Mendelian randomisation analyses have been performed to assess the effects of thyroid dysfunction on uterine cancer using genetic data from a limited number (n = 1,931) of cases from the UK Biobank, primarily composed of endometrial cancer patients [15]. No significant effects were observed for hypothyroidism, hyperthyroidism or T4 but TSH was found to be nominally associated with reduced cancer risk, inconsistent with the previous observational findings.” |
| 3 | **Objectives** | State specific objectives clearly, including pre-specified causal hypotheses (if any). State that MR is a method that, under specific assumptions, intends to estimate causal effects | 5-7 | “By leveraging the random assortment of germline variants, which remain unaffected by disease or environmental factors, the Mendelian randomisation approach enables the assessment of causality while mitigating biases arising from confounding and reverse causation [4].”  “In the current study, we aimed to provide clarity on the effects of thyroid dysfunction by using genetic data from a much larger number of endometrial cancer cases (n = 12,270) provided by the Endometrial Cancer Association Consortium (ECAC) [16]. These Mendelian randomisation analyses included genetic instruments for hypothyroidism, hyperthyroidism, TSH, T3 and T4. Considering that autoimmune thyroid disease, including Hashimoto’s thyroiditis (hypothyroidism) and Graves’ disease (hyperthyroidism), is responsible for ~90% of all thyroid diseases [17], we included these conditions in our analyses; in addition to thyroid peroxidase (TPO) antibody positivity, a marker for autoimmune thyroid disease. Furthermore, given the role of obesity in the development of thyroid dysfunction and endometrial cancer,” |
|  | **METHODS** |  |  |  |
| 4 | **Study design and data sources** | Present key elements of the study design early in the article. Consider including a table listing sources of data for all phases of the study. For each data source contributing to the analysis, describe the following: |  |  |
|  | a) | Setting: Describe the study design and the underlying population, if possible. Describe the setting, locations, and relevant dates, including periods of recruitment, exposure, follow-up, and data collection, when available. | 7 | “For Mendelian randomisation analyses, we sourced GWAS summary statistics from studies described in **Supplementary Table 1**. For endometrial cancer risk, data were derived from the largest GWAS meta-analysis of endometrial cancer risk [16].” |
|  | b) | Participants: Give the eligibility criteria, and the sources and methods of selection of participants. Report the sample size, and whether any power or sample size calculations were carried out prior to the main analysis | 7 and 10 | “To avoid potential bias from overlapping samples with other GWAS datasets used in the current study, UK Biobank samples had been removed, resulting in 12,270 endometrial cancer cases and 46,126 controls for the generation of GWAS summary statistics [18]. For secondary analyses using this dataset, we used summary statistics from GWAS of cases with either endometrioid (8,758 cases and 46,126 controls) or non-endometrioid histology (1,230 cases and 35,447 controls). Histological subtypes of endometrial cancer were confirmed based on pathology reports, and details have been described previously [16, 19].”  “We used the online Mendelian randomisation power calculator mRnd (https://shiny.cnsgenomics.com/mRnd/) [39] to calculate the power for Mendelian randomisation analyses to detect an association between one standard deviation (SD) change in the trait of interest under three scenarios reflecting weak, moderate, and strong effects with cut-offs set to OR > 1.1, OR > 1.2, and OR > 1.4, respectively. For the calculation of power for continuous outcomes, variances of the exposure and outcome variables were both set to 1 given GWAS for outcome were conducted on the inverse-normalised variable.” |
|  | c) | Describe measurement, quality control and selection of genetic variants | 9-10 | “GWAS summary statistics from all phenotypes were used to identify genetic variants associated with exposures at genome-wide significance levels (p < 5 × 10-8); no such variants were available for non-endometrioid endometrial cancer. To identify independent IVs, we used a window of 10 Mb and maximal linkage disequilibrium of r2=0.001 between instruments in PLINK [30]. The linkage disequilibrium reference used a random sample of 10,000 unrelated participants from the UK Biobank [31]. Palindromic variants (i.e. those with A/T or G/C alleles) with a minor allele frequency ≥ 0.42 were excluded to prevent errors due to strand ambiguity. Variants from the SH2B3 locus were also excluded as this is a pleiotropic locus and, through GWAS, has been associated with many traits including hypothyroidism [32], Hashimoto’s thyroiditis [24], and endometrial cancer [16]. To assess weak instrument bias, we calculated the F-statistic [33] for each instrumental variable and all IVs of an exposure using the formula F = R2 × (N-1-k) / ((1-R2) × k), where R2 is the proportion of variance explained by the instrumental variable, N is the sample size, and k is the number of IVs included in the analysis [34]. For the calculation of F-statistic for each instrumental variable, k equals to 1. The proportion of variance explained by IVs was calculated using the formula in the Supplementary Methods of [34]. An F-statistic > 10 is often used to indicate sufficient instrument strength.” |
|  | d) | For each exposure, outcome, and other relevant variables, describe methods of assessment and diagnostic criteria for diseases | Supplementary Table 1 | This information can be found in the publications describing the cohorts used to derive exposure and outcome data. |
|  | e) | Provide details of ethics committee approval and participant informed consent, if relevant | Supplementary Table 1 | This information can be found in the publications describing the cohorts used to derive exposure and outcome data. |
| 5 | **Assumptions** | Explicitly state the three core IV assumptions for the main analysis (relevance, independence and exclusion restriction) as well assumptions for any additional or sensitivity analysis | 10 and 11 | “The IVW method uses all available IVs and thus has the most power to detect an association assuming that the IVs meet certain criteria: 1) they are reliably associated with the exposure of interest; 2) they are not associated with any confounders that mediate the association between exposure and outcome; and 3) they only affect the outcome through the exposure, without any horizontal pleiotropy.”  “The MR-Egger method relaxes assumption 2) by allowing the intercept of the regression to be non-zero [40]. Even when there this assumption is violated for all IVs, this method still returns an unbiased causal estimate if the magnitude of the horizontal pleiotropic effects are independent of the variant-exposure effects. The MR-Egger method also provides a formal test of the existence of horizontal pleiotropy. The weighted median method assumes at least half of the variants are valid IVs [41].” |
| 6 | **Statistical methods: main analysis** | Describe statistical methods and statistics used |  |  |
|  | a) | Describe how quantitative variables were handled in the analyses (i.e., scale, units, model) | 9 | For quantitative traits, such as circulating thyroid hormones (i.e., TSH, T3, or T4), IVs are reported as the association with one standard deviation increase in the levels of the respective hormone. |
|  | b) | Describe how genetic variants were handled in the analyses and, if applicable, how their weights were selected | 10-11 | “For free T3, the only exposure with a single IV, we used the Wald ratio to estimate effects. For all other exposures, multiple IVs were combined using the inverse-variance weighted (IVW) random effect method as the primary analysis.” |
|  | c) | Describe the MR estimator (e.g. two-stage least squares, Wald ratio) and related statistics. Detail the included covariates and, in case of two-sample MR, whether the same covariate set was used for adjustment in the two samples | 9 | The MR estimators are described above in 6b. The covariate sets vary between GWAS – details can be found in the corresponding publications (Supplementary Table 1). |
|  | d) | Explain how missing data were addressed | 11 | “We were unable to study free T3 or TPO antibody positivity as outcomes due to the unavailability of full GWAS summary statistics for these phenotypes.” |
|  | e) | If applicable, indicate how multiple testing was addressed | 10 | “To determine the effects of thyroid dysfunction phenotypes on endometrial cancer risk, we used a Bonferroni correction threshold for significance, adjusting for the testing of 11 traits (p < 4.5 × 10-3).” |
| 7 | **Assessment of assumptions** | Describe any methods or prior knowledge used to assess the assumptions or justify their validity | 10 | “To detect violations in the underlying Mendelian randomisation assumptions, we assessed heterogeneity in the causal effect estimates across IVs by calculating Cochran’s Q statistic and its associated p-value.].” |
| 8 | **Sensitivity analyses and additional analyses** | Describe any sensitivity analyses or additional analyses performed (e.g. comparison of effect estimates from different approaches, independent replication, bias analytic techniques, validation of instruments, simulations) | 10-12 | “We also performed bidirectional analyses, using thyroid dysfunction phenotypes as outcomes and risk of endometrial cancer (and its histological subtypes) as exposures. To investigate the involvement of BMI in potential causal relationships, we conducted bidirectional MR analysis between BMI and exposures that were significantly associated with endometrial cancer risk.”  “Therefore, to account for potential violations of these assumptions and evaluate the robustness of significant associations identified by IVW analysis, we conducted sensitivity analyses using the MR-Egger, weighted median and the Mendelian Randomisation pleiotropy residual sum and outlier (MR-PRESSO) methods. The MR-Egger method relaxes assumption (2) by allowing a non-zero regression intercept [41]. This provides an unbiased causal estimate even when assumption (2) is violated for all IVs, as long as the magnitude of the horizontal pleiotropic effects is independent of the variant-exposure effects. The MR-Egger method also provides a formal test of the existence of horizontal pleiotropy [41]. The weighted median method assumes at least half of the variants are valid IVs and so provides a reliable estimate even if the assumptions for some IVs are violated [42]. Lastly, MR-PRESSO identifies and adjusts for outlier IVs displaying horizontal pleiotropic effects [43].  “To investigate the independent effects of BMI and hypothyroidism on endometrial cancer risk, we conducted multivariable Mendelian randomisation analysis. We first clumped genetic variants with GWAS summary statistics for BMI and hypothyroidism together in PLINK using a window of 10 Mb and a maximal linkage disequilibrium of r2=0.001 between variants, as per the univariable Mendelian randomisation analyses. As there are far more genetic variants associated with BMI, we preferentially selected variants for hypothyroidism that were in linkage disequilibrium with variants associated with BMI to maximise the number of IVs for hypothyroidism. A list of the selected IVs are provided in Supplementary Table S4. The multivariable analysis was performed using the Mendelian Randomisation (version 0.6.0) R package [43]. R code for the multivariable analysis can be found in Supplementary Information. To confirm the robustness of the results from the multivariable analysis, we also reran univariable Mendelian randomisation analyses with endometrial cancer risk as the outcome using only IVs included the multivariable analysis for BMI or hypothyroidism risk.” |
| 9 | **Software and pre-registration** |  |  |  |
|  | a) | Name statistical software and package(s), including version and settings used | 10-12 | Software are described in 6(b) and 8 above. |
|  | b) | State whether the study protocol and details were pre-registered (as well as when and where) | N/A | This is a secondary analysis based on summary statistics from existing, published studies. The ethical approval and informed consent have been obtained by all original studies. |
|  | **RESULTS** |  |  |  |
| 10 | **Descriptive data** |  |  |  |
|  | a) | Report the numbers of individuals at each stage of included studies and reasons for exclusion. Consider use of a flow diagram | 7 | See 4(b). |
|  | b) | Report summary statistics for phenotypic exposure(s), outcome(s), and other relevant variables (e.g. means, SDs, proportions) | 7-8 | Detailed information can be found in the corresponding GWAS publications in Supplementary Table 1. |
|  | c) | If the data sources include meta-analyses of previous studies, provide the assessments of heterogeneity across these studies | 7-8 | Detailed information can be found in the corresponding GWAS publications in Supplementary Table 1. |
|  | d) | For two-sample MR:  i.  Provide justification of the similarity of the genetic variant-exposure associations between the exposure and outcome samples  ii.  Provide information on the number of individuals who overlap between the exposure and outcome studies | 7-8 | Except for Graves’ disease, GWAS data were primarily derived from European populations, “For endometrial cancer risk, data were derived from the largest GWAS meta-analysis of endometrial cancer risk [16]. To avoid potential bias from overlapping samples with other GWAS datasets used in the current study, UK Biobank samples had been removed, resulting in 12,270 endometrial cancer cases and 46,126 controls for the generation of GWAS summary statistics [18].” |
| 11 | **Main results** |  |  |  |
|  | a) | Report the associations between genetic variant and exposure, and between genetic variant and outcome, preferably on an interpretable scale | Supplementary Table 2 |  |
|  | b) | Report MR estimates of the relationship between exposure and outcome, and the measures of uncertainty from the MR analysis, on an interpretable scale, such as odds ratio or relative risk per SD difference | 12-13 and Figure 1 | “Mendelian Randomisation analysis revealed a significant causal association between hypothyroidism and decreased endometrial cancer risk (OR = 0.93; 95% CI 0.89-0.97; p = 3.96 × 10-4; Figure 1A). Although no other thyroid dysfunction phenotype passed the Bonferroni multiple correction threshold for association with endometrial cancer risk (p < 4.5 × 10-3; Figure 1), we did observe a suggestive causal association between Hashimoto’s thyroiditis and decreased endometrial cancer risk (OR = 0.92, 95% CI 0.86-0.99; p = 0.03; Figure 1C).” |
|  | c) | If relevant, consider translating estimates of relative risk into absolute risk for a meaningful time period | N/A |  |
|  | d) | Consider plots to visualize results (e.g. forest plot, scatterplot of associations between genetic variants and outcome versus between genetic variants and exposure) | Figure 1 |  |
| 12 | **Assessment of assumptions** |  |  |  |
|  | a) | Report the assessment of the validity of the assumptions | 13 and Supplementary Table 5 | “In analysis of endometrial cancer histological subtypes, we also found a significant causal association between hypothyroidism and decreased endometrioid endometrial cancer risk (OR = 0.93, 95% CI 0.88-0.98; p = 4.02 × 10-3; Figure 1A). However, there was some evidence of IV heterogeneity in this analysis from Cochran’s Q statistic (p < 0.05; Supplementary Table 5).” |
|  | b) | Report any additional statistics (e.g., assessments of heterogeneity across genetic variants, such as *I^2^*, Q statistic or E-value) | Supplementary Table 2, 5 |  |
| 13 | **Sensitivity analyses and additional analyses** |  |  |  |
|  | a) | Report any sensitivity analyses to assess the robustness of the main results to violations of the assumptions | 13-14 | “We performed sensitivity analyses to evaluate the associations of hypothyroidism and Hashimoto’s disease with endometrial cancer risk. These analyses supported the association between hypothyroidism and decreased risk of endometrial cancer, as demonstrated by significant associations in the weighted median and MR-PRESSO analyses (Figure 2A). For the association of hypothyroidism with endometrioid endometrial cancer risk, the MR-Egger analysis showed a discordant direction of effect (Figure 2B) and suggested the presence of horizontal pleiotropy (Supplementary Table 5). However, by removing an IV that contributed to heterogeneity (Supplementary Table 5), the MR-PRESSO analysis demonstrated a significant association between hypothyroidism and decreased endometrioid endometrial cancer risk, consistent with the IVW analysis (Figure 2B). Lastly, the MR-Egger analysis again provided a discordant direction of effect for the association of Hashimoto’s thyroiditis with endometrial cancer risk (Figure 2C), but there was no evidence of pleiotropy (Supplementary Table 5). Furthermore, the directionality of the MR-PRESSO and weighted median analyses were consistent with the association of Hashimoto’s thyroiditis with reduced endometrial cancer risk and (Figure 2C).” |
|  | b) | Report results from other sensitivity analyses or additional analyses | 14 | “To assess the effect of BMI on the relationship between hypothyroidism and endometrial cancer risk, we firstly performed bidirectional Mendelian randomisation analysis of BMI and hypothyroidism. Our findings revealed that increased BMI was very strongly associated with increased hypothyroidism risk but there was no evidence for a significant effect of predisposition to hypothyroidism on BMI (Supplementary Table 7). Although some IV heterogeneity was detected in the IVW analysis of BMI’s effect on hypothyroidism, the finding remained significant in all sensitivity analyses (Supplementary Table 7). We next performed multivariable Mendelian randomisation, which demonstrated that both BMI and hypothyroidism independently associated with endometrial cancer risk (Figure 3A-B). Similar associations were also observed for endometrioid endometrial cancer risk (Figure 3A-B). Notably, the multivariable analyses showed slightly stronger associations compared to the univariable analyses, particularly for the effect of BMI on endometrial cancer risk, including the histological subtypes (Figure 3B). These observations suggest that hypothyroidism acts as a mediator in the causal pathway between BMI and endometrial cancer (Figure 3C). Specifically, increased BMI raises the risk of hypothyroidism, which in turn has a protective effect on endometrial cancer.” |
|  | c) | Report any assessment of direction of causal relationship (e.g., bidirectional MR) | 13 | “Bidirectional Mendelian randomisation analysis did not support effects of endometrial cancer risk on thyroid dysfunction phenotypes (Supplementary Table 6).” |
|  | d) | When relevant, report and compare with estimates from non-MR analyses | N/A |  |
|  | e) | Consider additional plots to visualize results (e.g., leave-one-out analyses) | Figures 2 and 3 |  |
|  | **DISCUSSION** |  |  |  |
| 14 | **Key results** | Summarize key results with reference to study objectives | 18 | “Our study provides clarity into the relationship between thyroid dysfunction and endometrial cancer risk. Mendelian Randomisation analyses provide evidence of a causal association between hypothyroidism and decreased risk of endometrial cancer, particularly the endometrioid subtype. Notably, we did not find any significant associations between thyroid hormone levels and endometrial cancer risk. The underlying mechanism for the potential protective effect of hypothyroidism is unclear and further study is needed, but autoimmunity, driven by Hashimoto's disease, may play a role. Multivariable analyses revealed that BMI and hypothyroidism independently influence endometrial cancer risk, with hypothyroidism appearing to act as a mediator and attenuating the effect of BMI.” |
| 15 | **Limitations** | Discuss limitations of the study, taking into account the validity of the IV assumptions, other sources of potential bias, and imprecision. Discuss both direction and magnitude of any potential bias and any efforts to address them | 17 | “Several limitations should be acknowledged when interpreting our results. Firstly, the statistical power of our analyses varied across thyroid dysfunction phenotypes, with limited power to detect low to moderate effects of phenotypes such as hyperthyroidism, Graves' disease, and TPO antibody positivity. As a result, the lack of significant associations for these phenotypes could be the result of insufficient statistical power rather than a true absence of association. This limitation highlights the need for larger GWAS datasets and additional IVs to capture more of the trait variance for future analyses. Secondly, the analyses of the non-endometrioid subtype were exploratory in nature and should also be interpreted with caution. There was low statistical power to detect associations with this outcome and significant heterogeneity exists within the non-endometrioid subtype, which encompasses tumours with serous, clear cell, and other histologies.” |
| 16 | **Interpretation** |  |  |  |
|  | a) | Meaning: Give a cautious overall interpretation of results in the context of their limitations and in comparison with other studies | 15-16 | See 15 above and also “We did not detect any significant associations between thyroid hormone levels and endometrial cancer risk in our analysis. This contrasts with a previous Mendelian randomisation study by Yuan et al., which reported a nominal association between TSH levels and uterine cancer risk [15]. Yuan et al. also studied 21 other site-specific cancers and found that TSH levels were associated with reduced risk of overall cancer, with individual associations for reduced risk of breast and thyroid cancer. Despite having sufficient statistical power to detect weak to moderate effects of TSH levels, we found no evidence of such an association with endometrial cancer risk. However, Yuan et al. also identified a causal association between hypothyroidism and decreased thyroid cancer risk, consistent with our findings.” |
|  | b) | Mechanism: Discuss underlying biological mechanisms that could drive a potential causal relationship between the investigated exposure and the outcome, and whether the gene-environment equivalence assumption is reasonable. Use causal language carefully, clarifying that IV estimates may provide causal effects only under certain assumptions | 16 | “The mechanism underlying the potential protective effect of hypothyroidism on endometrial cancer remains unclear. It is noteworthy that hypothyroidism often develops in endometrial cancer patients as a result of immune suppression therapy [46, 47]. This observation suggests that the effect of hypothyroidism on endometrial cancer may be linked to autoimmunity, driven by Hashimoto's disease. Indeed, systemic lupus erythematosus (SLE), another autoimmune disease, has been robustly associated with reduced endometrial cancer risk in Mendelian randomisation analyses [48]. Furthermore, Mendelian randomisation analysis indicates that there is bidirectional relationship between hypothyroidism and SLE, with a particularly strong effect of hypothyroidism on increased SLE risk [49]. The protective effect of SLE on endometrial cancer has been proposed to be related to antinuclear autoantibodies that target cancer cells with defective DNA repair [48]. Thus, thyroid autoantibodies may have a similar effect. Supporting this hypothesis, an observational study found that uterine cancer was significantly less prevalent in thyroid disease patients who had thyroid autoantibodies compared to thyroid disease patients who tested negative for autoantibodies [8].” |
|  | c) | Clinical relevance: Discuss whether the results have clinical or public policy relevance, and to what extent they inform effect sizes of possible interventions | N/A |  |
| 17 | **Generalizability** | Discuss the generalizability of the study results (a) to other populations, (b) across other exposure periods/timings, and (c) across other levels of exposure | 17 | “Finally, the generalizability of our findings may be limited to populations of European ancestry, as the genetic data used in our study predominantly represent individuals from this genetic background. To validate and generalize these findings, it is crucial for future studies to encompass more diverse populations.” |
|  | **OTHER INFORMATION** |  |  |  |
| 18 | **Funding** | Describe sources of funding and the role of funders in the present study and, if applicable, sources of funding for the databases and original study or studies on which the present study is based | 18 | This research was funded by an Investigator grant from the National Health and Research Council of Australia, grant number APP1173170, and a project grant co-funded by Worldwide Cancer Research and Cancer Australia, grant number 22-0253. |
| 19 | **Data and data sharing** | Provide the data used to perform all analyses or report where and how the data can be accessed, and reference these sources in the article. Provide the statistical code needed to reproduce the results in the article, or report whether the code is publicly accessible and if so, where | 19 | All data analysed in this study are publicly available and their sources have been referenced throughout the manuscript and supplementary materials. |
| 20 | **Conflicts of Interest** | All authors should declare all potential conflicts of interest | 18 | The authors declare that they have no competing interests. |

This checklist is copyrighted by the Equator Network under the Creative Commons Attribution 3.0 Unported (CC BY 3.0) license.

1.	Skrivankova VW, Richmond RC, Woolf BAR, Yarmolinsky J, Davies NM, Swanson SA, et al. Strengthening the Reporting of Observational Studies in Epidemiology using Mendelian Randomization (STROBE-MR) Statement. JAMA. 2021;under review.

2.	Skrivankova VW, Richmond RC, Woolf BAR, Davies NM, Swanson SA, VanderWeele TJ, et al. Strengthening the Reporting of Observational Studies in Epidemiology using Mendelian Randomisation (STROBE-MR): Explanation and Elaboration. BMJ. 2021;375:n2233.
